# AI-Enhanced Integration of Genetic and Medical Imaging Data for Risk Assessment of Type 2 Diabetes

**DOI:** 10.1101/2023.08.14.23294093

**Authors:** Yi-Jia Huang, Chun-houh Chen, Hsin-Chou Yang

**Affiliations:** Institute of Public Health, National Yang-Ming Chiao-Tung University, Taipei, Taiwan; Institute of Statistical Science, Academia Sinica, Taipei, Taiwan

**Keywords:** Type 2 diabetes (T2D), risk assessment, Taiwan Biobank, single nucleotide polymorphism (SNP), medical imaging, polygenic risk score (PRS), eXtreme Gradient Boosting (XGBoost)

## Abstract

Type 2 diabetes (T2D) is a global public health concern due to its increasing prevalence. Risk assessment and early detection of T2D are vital in improving individuals’ health, reducing the burden on health insurance, and enhancing well-being. This study leverages artificial intelligence (AI), specifically eXtreme Gradient Boosting (XGBoost), to develop predictive models for T2D based on genetic and medical imaging data. The study aims to establish a prediction model and identify high-risk subgroups for T2D within a cohort of 68,911 Taiwan Biobank (TWB) participants. The approach integrates the Polygenic Risk Score (PRS) and Multi-image Risk Score (MRS) with demographic factors and environmental exposures to assess T2D risk. The model’s performance is evaluated using the Area Under the Receiver Operating Curve (AUC). Results demonstrate that genetic information alone is insufficient for accurate T2D prediction (AUC = 0.73), whereas medical imaging data, including abdominal ultrasonography, vertebral artery ultrasonography, bone density scan, and electrocardiography, significantly improves prediction accuracy (AUC = 0.89). The best-performing model integrates genetic, medical imaging, and demographic variables (AUC = 0.94), successfully identifying subgroups at high risk of developing T2D. The study also presents an online risk assessment website for T2D. In summary, this research represents the first integration of genetic and medical imaging data for T2D risk assessment. The genetic-only model outperforms previous genetic prediction studies, and integrating genetic and medical imaging information significantly enhances prediction. By utilizing artificial intelligence to analyze genetic, medical imaging, and demographic factors, this study contributes to early detection and precision health of T2D.

## Introduction

Type 2 diabetes (T2D) is a prevalent global health concern, comprising almost 90% of diabetes mellitus (DM) cases ^1^. T2D is associated with severe complications such as retinopathy, nephropathy, and cardiovascular diseases, significantly impacting health and quality of life and increasing healthcare expenses ^2^. Early detection and risk assessment of T2D are crucial for effective health management. T2D has a global prevalence of 6% ^3^. In Taiwan, the prevalence is even higher, at approximately 10%. The mortality and economic burden in medical care among T2D patients increase significantly over time ^4^. T2D has a polygenic and multifactorial mode of inheritance ^5,6^. The significant risk factors include genetic components, food intake, and environmental exposures ^7,8^.

Genome-wide association studies (GWAS) have identified T2D susceptibility loci and genes, which have been used to develop T2D prediction models ^9-11^. Polygenetic risk scores (PRS) and weighted PRS have attracted attention for the genetic prediction of T2D ^12-14^. However, the prediction accuracy must be elevated for clinical use ^15^. Recent studies have combined single nucleotide polymorphisms (SNPs) from multi-ethnic GWAS to calculate PRS and improve prediction accuracy ^16,17^. Methods, such as PRS-CSx, have been developed to integrate GWAS summary statistics from multiple ethnic groups and combine multiple PRSs with weights that consider linkage disequilibrium ^18-20^. The use of PRS for T2D risk assessment and prediction is crucial in clinical application and precision medicine ^21^.

Recent smart medicine and precision health studies have highlighted the utility of medical imaging analysis in disease diagnosis and prediction, in addition to genetic markers. Moreover, previous research has demonstrated the association of several diseases with T2D ^22,23^, some of which can be diagnosed using medical imaging techniques. For instance, nonalcoholic fatty liver can be diagnosed through abdominal (ABD) ultrasonography ^24^, osteoporosis through bone mineral density (BMD) ^25^, and cardiovascular disease through electrocardiography (EKG) ^26^. These T2D-associated diseases can be effectively diagnosed and detected using medical imaging analysis. Considering this, our study incorporates genetic markers and medical imaging data to assess the risk of T2D. This approach enables a comprehensive evaluation and potential improvement in T2D prediction and risk assessment.

Artificial intelligence, which encompasses machine learning and deep learning, has found extensive applications in genetic research, including disease diagnosis, classification, and prediction using supervised learning ^27,28^. Extreme Gradient Boosting (XGBoost), a supervised tree-based machine learning approach ^29^, has demonstrated superior performance in classification and prediction. Successful applications of XGBoost in precision medicine include chronic kidney disease diagnosis ^30^, orthopedic auxiliary classification ^31^, chronic obstructive pulmonary prediction ^32^, and multiple phenotypes prediction ^33^.

Taiwan Biobank (TWB), established in 2012, is a valuable resource for the integrative analysis of genetic and medical imaging data ^34^. The TWB enrolled participants aged over 20 and collected baseline questionnaires, blood, urine samples, and their biomarkers of lab tests, and genotyping data from all participants. Follow-up data, including repeated questionnaires, biomarker measurements, and medical imaging data, were collected every two to four years. Medical imaging data includes ABD, vertebral artery ultrasonography (VAU), BMD, EKG, and thyroid ultrasonography (TU). The integrative analysis of genetic and medical imaging data holds great promise for disease risk assessment and prediction, as demonstrated by recent studies ^35-38^. Here, we present the first study integrating genome-wide SNPs and multimodality imaging data from the TWB for T2D risk assessment. We developed machine learning models incorporating genetic information, medical imaging, demographic variables, and other risk factors. Furthermore, we identified high-risk subgroups for T2D, providing insights into T2D precision medicine.

## Study participants and variables

This study included a genetic-centric analysis (Analysis 1) and genetic-imaging integrative analysis (Analysis 2). A total of 68,911 participants in the TWB were analyzed.

In the genetic-centric analysis, 50,984 participants who had only baseline data (i.e., without follow-up data) were used as the training and validation samples; they consisted of 2,531 self-reported T2D patients and 48,453 self-reported non-T2D controls (**Fig. 1A** and **Fig. S1A**). Here, 9,763 participants who had both baseline and follow-up data were used as the testing samples, where 8,827 and 936 participants were recruited as the first and second testing datasets; they consisted of 528 self-reported T2D patients and 9,235 self-reported non-T2D controls at baseline; 767 self-reported T2D patients and 8,996 self-reported non-T2D controls at follow-up (**Fig. 1A** and **Fig. S1A**).

**Figure 1.**
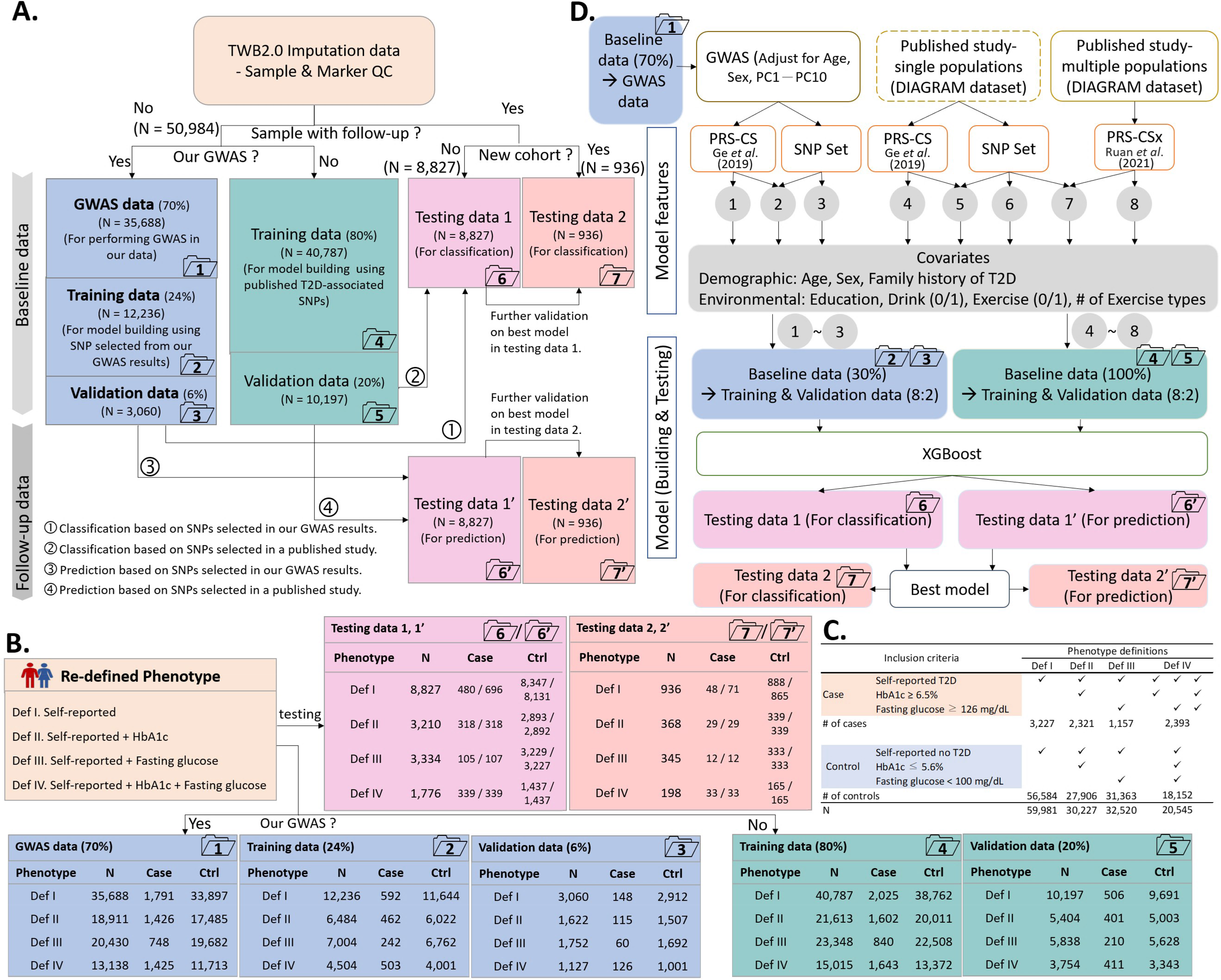
Flowchart of genetic-centric analysis. **(A) Data partitioning.** The dataset containing information from 60,747 individuals was divided into several subsets: (i) The GWAS samples (Dataset 1, N = 35,688), training samples (Dataset 2, N = 12,236; Dataset 4, N = 40,787), and validation samples (Dataset 3, N = 3,060; Dataset 5, N = 10,197). For classification analysis, testing samples comprised Dataset 6 (N = 8,827) and Dataset 7 (N = 936), while for prediction analysis, they were represented as Datasets 6’ (N = 8,827) and Dataset 7’ (N = 936); **(B) Sample size.** Total sample size, along with the number of cases and the number of controls, are shown for each of the four phenotype definitions in Datasets 1 – 7; **(C) Phenotype definition criteria.** The definition and sample size for the four T2D phenotype definitions is shown. **(D) Analysis flowchart.** The analysis flow comprises three steps, starting with selecting T2D-associated SNPs and PRS, then selecting demographic and environmental covariates, and the best XGBoost model was established using the selected features.

In addition to the self-reported T2D, hemoglobin A1C (HbA1c) and fasting glucose (GLU-AC) collected in both baseline and follow-up were used to refine the self-reported T2D phenotype (**Figs. 1B** and **1C**). Other variables in the genetic-centric analysis (**Fig. 1D**) are illustrated as follows: Demographic variables included age, sex, and family history of T2D. Four types of family history were: T2D occurrence in parents (Yes or No), in sibs (Yes or No), in any of parents and sibs (Yes or No), and in father, mother, brother(s), and sister(s) (0, 1, 2, 3, or 4). Environmental exposures included education level, drinking level, exercise habits, and the number of exercise types.

Whole-genome genotyping using one of two SNP arrays was performed based on the samples in the baseline. TWBv1.0 SNP array with approximately 650,000 SNP markers or TWBv2.0 SNP array with approximately 750,000 SNP markers was employed. Imputation was performed based on the 1KG-EAS panel ^39^. The SNPs with an info score of less than 0.9 were removed ^40^. Sample and marker quality controls followed the procedures of Yang et al. ^41^. External information about T2D-associated SNP sets and effect sizes based on the GWAS summary statistics of T2D were collected (**Supplemental Text 1**).

In the genetic-imaging integrative analysis, 17,785 participants who had both genetic data and medical imaging data were analyzed (**Fig. 2A** and **Fig. S1B**); they consisted of 1,366 self-reported T2D patients and 16,419 self-reported non-T2D controls (**Fig. 2A**); here, the case and control were defined based on the questionnaire at follow-up rather than baseline. For example, based on the T2D Definition IV (**Fig. 1C**), 7,786 participants, which consisted of 1,118 cases and 6,668 controls, were analyzed (**Fig. 2A**). Imaging report variables in the genetic-imaging integrative analysis (Upper left in **Fig. 2A**) consisted of 28 ABD features, 29 VAS features, 85 BMD features, and 10 EKG features (**Supplemental Table S1**). TU features were not included in our analysis because of a small sample size. The details about the medical imaging protocol can be referred to TWB (https://www.biobank.org.tw/about_value.php). In the flowchart of PRS calculation, external information about T2D-associated SNP sets and GWAS summary statistics from DIAGRAM ^42^ are provided (**Fig. 2B**).

**Figure 2.**
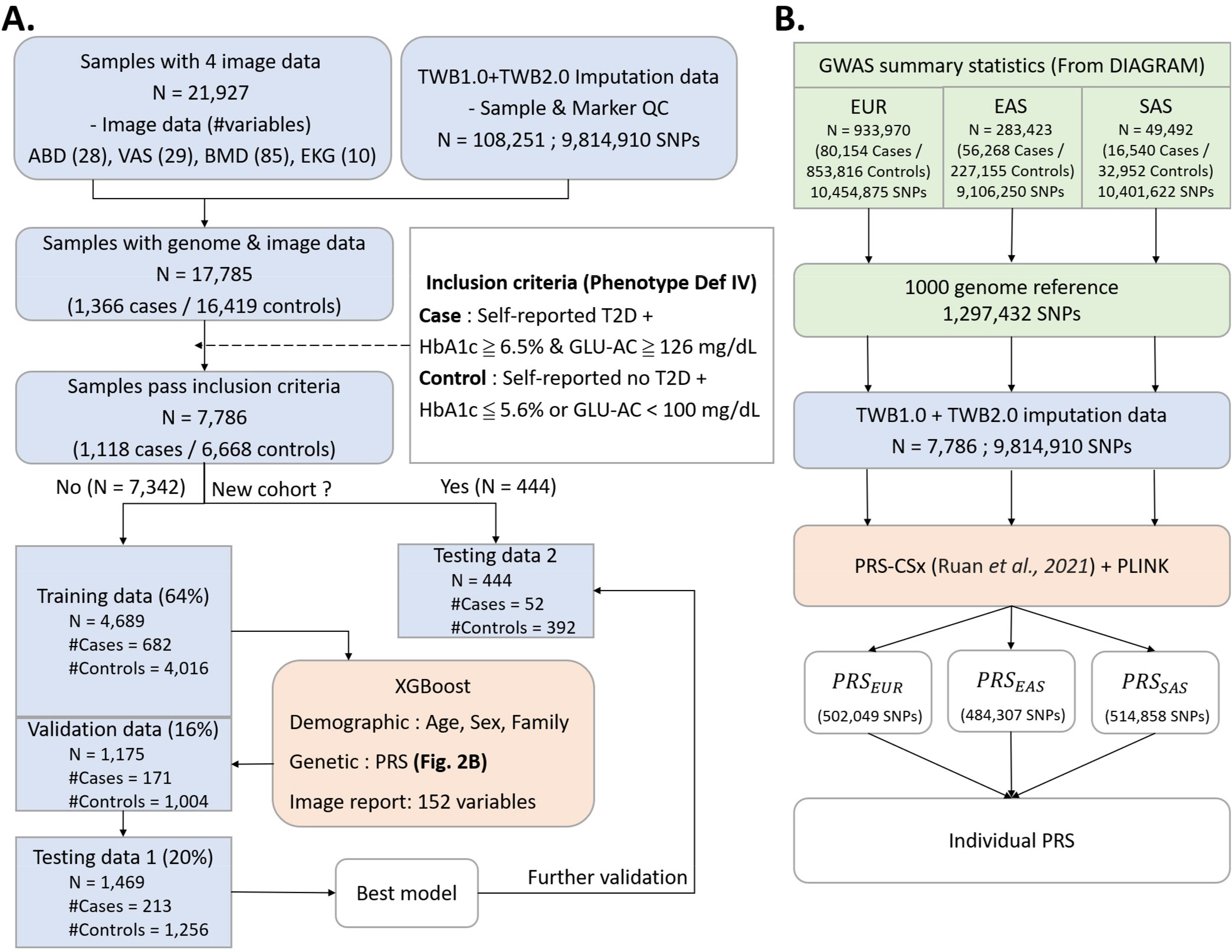
Flowchart of genetic-image integrative analysis. **(A) Data partitioning and model training.** To illustrate the process, Phenotype Definition IV was used as an example. The data containing information from 7,786 individuals were divided into four subsets: a training dataset (N = 4,689), a validation dataset (N = 1,175), and two independent testing datasets (N = 1,469 for the first dataset and N = 444 for the second independent dataset). Subsequently, the best XGBoost model was established. **(B) Flowchart of PRS construction.** The construction of PRS was carried out using PRS-CSx based on the GWAS summary statistics obtained from the analysis of the DIAGRAM Project.

## Methods

### Classification and prediction for T2D

XGBoost algorithm ^29^ was employed to classify and predict T2D using the *XGBClassifier* function in the Python package *xgboost* based on a set of features encompassing genetic, demographic, environmental exposures, and imaging report variables. Both classification and prediction models were trained and validated based on the baseline data (Datasets 1 – 5 in **Fig. 1A**). Final classification models were built and tested based on the baseline phenotype data (Dataset 6 in **Fig. 1A**) and further replicated based on the second independent testing dataset (Dataset 7 in **Fig. 1A**). Final prediction models were built and tested based on the follow-up phenotype data (Dataset 6’ in **Fig. 1A**) and replicated based on the second independent testing dataset (Dataset 7’ in **Fig. 1A**). The illustration of the data used for classification and prediction tasks in genetic-centric analysis (Analysis 1) and genetic-imaging integrative analysis (Analysis 2) are provided in **Supplemental Table S2**.

The XGBoost models were trained with the following default parameter settings: maximum depth equal to 6, learning rate equal to 0.3, the value of the regularization parameter alpha (L1) was set to 0, and lambda (L2) was set as 1, the number of boosting stages was 100, and the early-stop parameter was set to 30.

The area under the receiver operating curve (AUC) was calculated to evaluate the model’s overall performance. The DeLong test (DeLong et al., 1988) examined the difference between AUCs using the *roc.test* function in the R package *pROC*. Bonferroni’s correction ^43^ was applied to control for a family-wise error rate in multiple comparisons. Accuracy, sensitivity, specificity, and F1-score were calculated to evaluate the performance of the established models, where the optimal cut-off value of the XGBoost model was calculated using the Youden index ^44^ in the validation data.

### Event history analysis and online risk assessment

In the genetic-centric analysis, multivariate Cox regression ^45^ was employed using the *cox.ph* function in the R package *survival* to identify important risk factors for the T2D event time and estimate the T2D-free probability in the testing datasets. The event was defined as the occurrence of T2D in the follow-up for non-T2D participants at baseline, and the event time was calculated by the duration from the baseline to the follow-up. Median event time in weeks was also calculated. Because medical imaging data were only available in the follow-up, the genetic-imaging integrative analysis applied multivariate logistic regression ^46^ using the R *glm* function to identify important risk factors for T2D events and estimate the T2D-free probability in the testing datasets. To assess the impact of exercise on HbA1c, linear regression analysis using the R *glm* function was performed. In addition, we established a website at https://hcyang.stat.sinica.edu.tw/software/T2D_web/header.php to provide an online risk assessment for T2D.

## Results

### Genetic-centric analysis

#### Comparison of prediction models

We evaluated the prediction performance under different scenarios hierarchically (the best scenario at a previous variable was given for a discussion of the next variable) in the following order: the sources and significance levels of T2D-associated SNPs (**Fig. 3A** and **Fig. S2**), T2D phenotype definitions (**Fig. 3B**), family history variable combinations (**Fig. 3C** and **Fig. S3**), demographic variable combinations (**Fig. 3D**), demographic and genetic variable combinations (**Fig. 3E**), and SNP and PRS combinations (**Figs. 3F** and **3G**). The findings are summarized as follows: First, using T2D-associated SNPs from the previous large-sample-size GWAS ^11^ as predictors had the highest AUC of 0.557, but its AUC was not significantly higher than that used the SNPs identified by our smaller-sample-size GWAS under different thresholds of statistical significance (**Fig. 3A**), although our GWASs did identify some T2D-associated SNPs (**Fig. S4**). Second, phenotype defined by self-reported T2D with HbA1C ≥6.5% or fasting glucose ≥126 mg/dL (i.e., T2D Definition IV) had the highest AUC 0.640, and its AUC was significantly higher than the AUCs of the other three T2D definitions (**Fig. 3B**). Third, sibs’ disease history had a significantly higher AUC 0.732 than parents’ disease history with an AUC 0.670 (p = 0.009). Moreover, additive parent-and-sib disease history had the highest AUC of 0.758. Its AUC was significantly higher than parent-only (p < 0.001) (**Fig. 3C**). Fourth, a joint effect of age, sex, and additive parent-sib disease history had the highest AUC of 0.884. Its AUC was significantly higher than other demographic variable combinations, except for the combination of age and additive parent-sib disease history (**Fig. 3D**). Fifth, whatever SNPs were included or not, demographic and PRS combinations outperformed the models without incorporation of PRS (**Fig. 3E**), although genetic factors only improved up to 3% of AUC conditional on demographic factors (age, sex, and family history of T2D). Finally, given T2D-associated SNPs, AUC significantly increased if PRS was included (**Fig. 3F**); T2D-associated SNPs provided a limited additional effect if PRS was already included (**Fig. 3G**).

**Figure 3.**
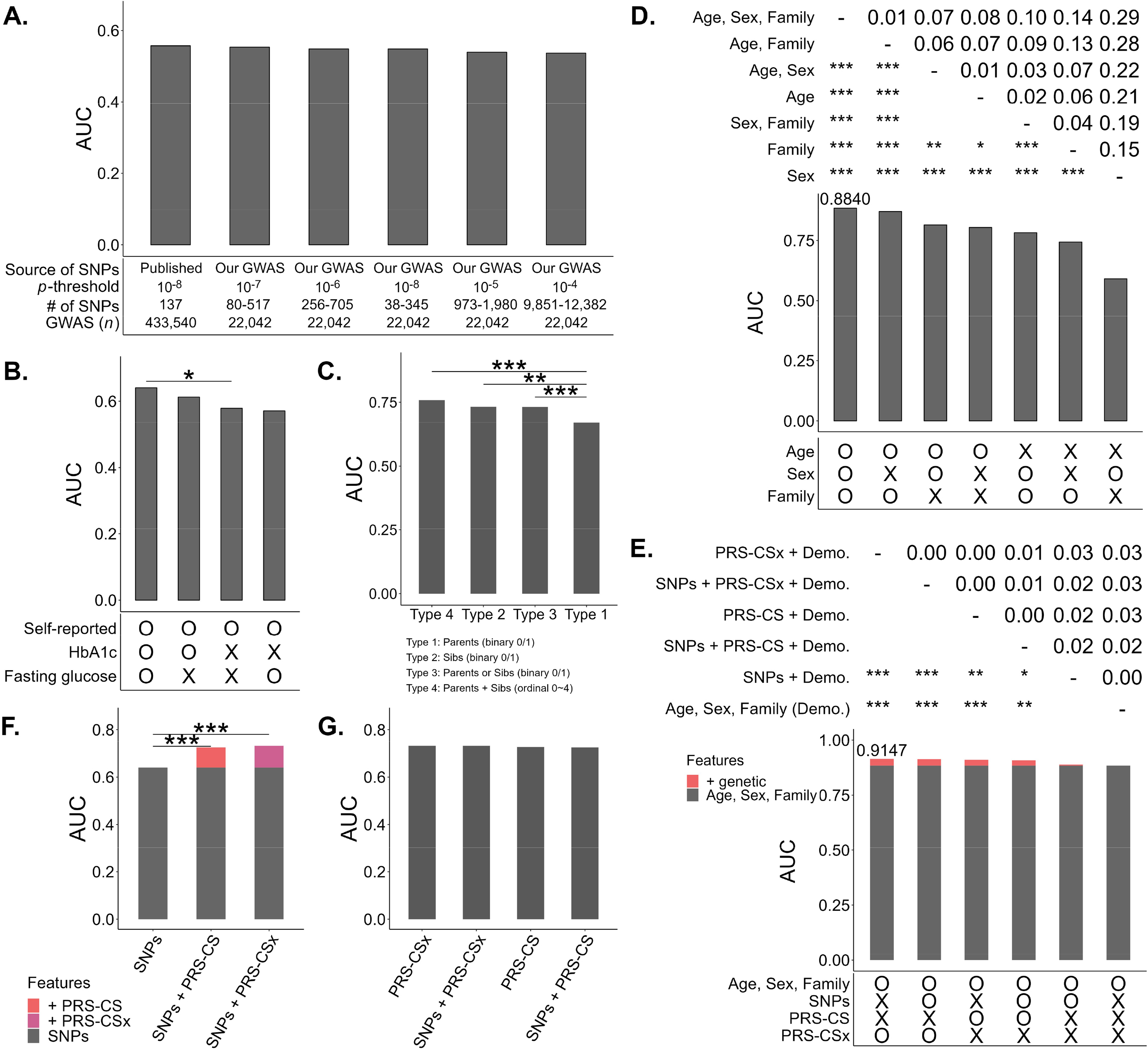
Model evaluation and comparison: Bar chart displaying AUC. **(A) SNP selection.** Model predictors were SNPs selected from published studies or our GWAS under different p-value thresholds. The average AUCs of prediction models for four phenotype definitions were compared. Incorporating SNPs from published GWAS with a large sample size has the highest AUC. **(B) T2D Phenotype Definition.** In addition to including the selected variables in Fig. 3A, the AUCs of four phenotype definitions were compared. T2D Phenotype Definition IV (i.e., phenotype defined by self-reported T2D, HbA1c, and fasting glucose) has the highest AUC. **(C) Family history of T2D.** In addition to including the selected variables in **Figs. 3A–3B**, the AUCs of the four types of T2D family history (i.e., (i): parents (binary factor), (ii) sibs (binary factor), (iii) either parents or sibs (binary factors), and (iv) both parents and sibs (ordinal factor)) were compared. Incorporating T2D family history that considers both parents’ and sibs’ disease history as ordinal predictors has the highest AUC. **(D) Demographic variables.** In addition to including the selected variables in **Figs. 3A–3C**, the AUCs of different combinations of demographic factors, including age, sex, and family history of T2D, are compared. Incorporating age, sex, and family history of T2D as predictors have the highest AUC. **(E) PRS and demographic variables.** In addition to including the selected variables in **Figs. 3A–3D**, the AUCs of different combinations of genetic variables, including SNPs, PRS-CS, and PRS-CSx, and demographic variables, including age, sex, and family history of T2D, are compared. Incorporating PRS and demographic factors as predictors have the highest AUC. **(F) Impact of including PRS after SNPs.** The AUCs of the models that consider SNPs, SNPs+PRS-CS, and SNPs+PRS-CSx as predictors are compared. PRS significantly improves prediction performance. PRS-CSx outperforms other genetic variables. **(G) Impact of including additional SNPs after PRS.** The AUCs of the models that consider additional SNPs given PRS in the model are compared. SNPs cannot further enhance prediction accuracy after PRS. PRS and PRS+SNP show similar prediction performances.

Among different prediction models, the model with predictors PRS-CSx, age, sex, and family history of T2D had the highest AUC 0.915 (**Fig. 4A**) for T2D Definition IV based on the first testing dataset (i.e., Dataset 6’ in **Fig. 1**). The optimal threshold, determined by the Youden index, for the fitted value that used to predict T2D or non-T2D in the XGboost model was 0.16. The Accuracy, Sensitivity, Specificity, and F1 indices were 0.843, 0.844, 0.843, and 0.672, respectively. Furthermore, the model was tested in the second independent testing dataset (i.e., Dataset 7’ in **Fig. 1**), and a promising result similar to the first testing dataset was found: AUC = 0.905, Accuracy = 0.843, Sensitivity = 0.846, Specificity = 0.842, and F1 = 0.644. AUCs are also provided for the other three T2D definitions (**Fig. S5**).

**Figure 4.**
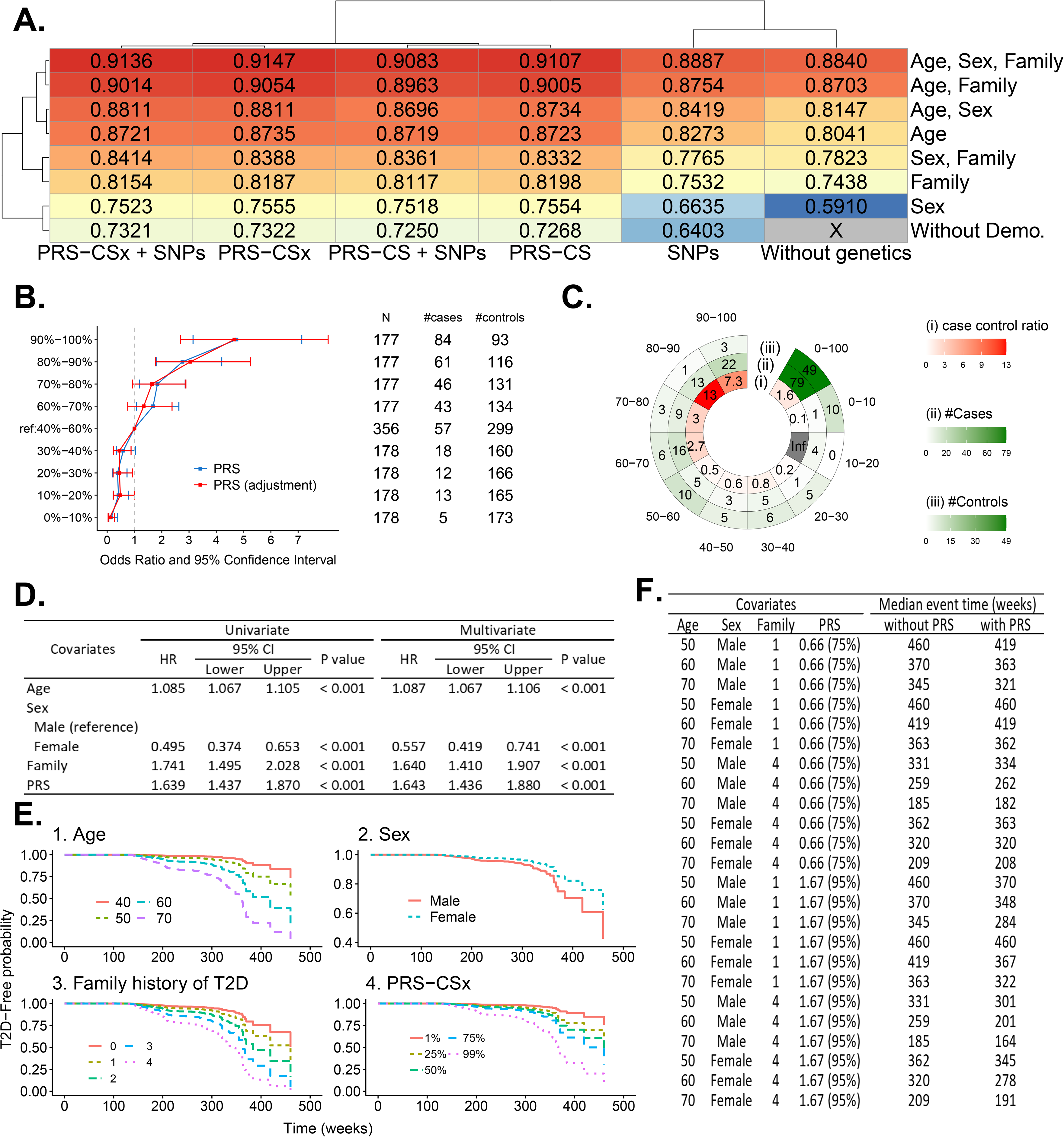
Results in the genetic-centric analysis. **(A) AUCs of all models based on Phenotype Definition IV.** A heatmap summarizes the AUCs of all models based on Phenotype Definition IV (i.e., T2D was defined by self-reported T2D, HbA1c, and fasting glucose). The genetic variables are shown on the X-axis, and the demographic variables are shown on the Y-axis. The best model comprises age, sex, family history of T2D, and PRS-CSx as predictors. **(B) Positive correlation between PRS and T2D odds ratio.** In each decile of PRS based on PRS-CSx, the odds ratio of T2D risk and its 95% confidence interval were calculated based on an unadjusted model (blue line) and an adjusted model considering age, sex, and T2D family history (red line). The reference group was the PRS group in the 40%–60% decile. **(C) High-risk group.** In the chart, the figures from the inner to the outer represent (i) the case-to-control ratio, (ii) the number of cases, and (iii) the number of controls in the PRS decile subgroups. A high-risk group was identified, comprising females older than 59 with a T2D family history and falling into the PRS group in the 80%–100% decile group. **(D) Association of age, sex, T2D family history, and PRS with T2D.** Hazard rates, 95% confidence intervals, and p-values based on univariate and multivariate Cox regression models demonstrate that age, sex, T2D family history, and PRS are significantly associated with T2D. **(E) Risk factors for T2D.** Predicted survival proportion curves reveal that elder persons, males, the number of parents and siblings who had T2D, and the high decile PRS group are risk factors for T2D risk. Age (older persons), sex (males), T2D family history (the number of parents and siblings who had T2D), and PRS (high decile PRS group) are risk factors (high-risk level) for T2D risk. **(F) Median event time of T2D.** Examples of the median even time for developing T2D are provided based on a multivariate Cox regression model, both without and with incorporating PRS. For instance, a male aged 50 with a T2D family history in one family member will develop T2D after 460 weeks, and the onset time is advanced to 419 weeks when PRS is further incorporated.

Further consideration of the environmental factors, including education level, drinking experience, exercise habits, the number of exercise types, and SNP-SNP interactions with and without SNPs’ main effect, had no improvement for T2D prediction (**Supplementary Table S3**). Considering model parsimony, these environmental factors and SNP-SNP interactions were not included in the final model. In addition to prediction models, classification models were also established. The AUCs in classification models (**Fig. S6**) were generally higher than those in prediction models (**Fig. S5**).

#### Assessment of PRS

The positive association between PRS and T2D risk is shown (**Fig. 4B**). Compared to the participants in the 40%–60% PRS decile group, those in the top 10% decile group had a 4.738-fold risk of developing T2D (95% confidence interval: 3.147–7.132, *p* < 0.001) and a 4.660-fold risk (95% confidence interval: 2.682–8.097, *p* < 0.001) after adjusting for age, sex, and family history. We identified a high-risk subgroup of women who were older than 59 and had a family history of T2D. The ratio of case vs. control sample size was as high as a 7.3–13.0-fold in the 80%–100% decile groups (**Fig. 4C**). The ratio was much higher than a 1.6-fold that did not consider PRS (i.e., PRS at 0%–100%) (**Fig. 4C**).

#### Risk of developing T2D

Among 8,347 non-T2D participants at baseline in the first testing dataset of 8,827 participants, 220 reported T2D in the follow-up. The duration from the baseline to the follow-up was treated as the event time. For the remaining 8,127 participants who reported having no T2D in the follow-up, their event time was censored. In multivariate Cox regression analysis, age, sex, family history of T2D, and PRS were all significantly associated with T2D (p<0.001) (**Fig. 4D**). Increased age, higher PRS, and stronger T2D family history had a higher T2D risk. The elderly male, with a strong family history and high PRS, had a severe T2D risk (**Fig. 4E** and **Fig. S7**). We also provided the predicted time-to-event (week) (**Fig. 4F**). For example, a 50-year-old man with one of his family members had T2D will achieve median T2D-free time after 460 weeks. The time to develop T2D was shortened to 419 weeks after considering a standardized PRS of 0.66 (equivalent to a PRS risk subgroup in the top 25% of the population).

To assess the impact of exercise on HbA1c, linear regression analysis was performed. It was observed that individuals engaging in regular exercise experienced a significant reduction in HbA1c by an average of 0.12% mg/dL (p = 0.023) compared to those who did not engage in regular exercise. Moreover, individuals with a high PRS who engaged in exercise demonstrated a greater reduction in HbA1c (0.13% mg/dL) than those with a low PRS (0.12% mg/dL). Additionally, individuals diagnosed with T2D experienced a noteworthy improvement of 0.3% mg/dL in HbA1c. In addition, among the various types of exercise, walking for fitness exhibited the most substantial effectiveness in reducing HbA1c (**Fig. S8**).

#### The ability of T2D early detection in our model

To investigate the early detection capability of our model for T2D, we performed an analysis focusing on 550 women participants older than 59 years, all of whom had a family history of T2D. We identified them as being at high risk if they possessed a high PRS, even though they were initially reported as non-T2D at baseline. Thirty-six of them changed to T2D, and 514 were still non-T2D at follow-up. We predicted their T2D status. G1 – G4 are the groups of participants in true positive, false negative, false positive, and true negative, respectively (**Fig. 5A**). We evaluated that G3 was indeed misclassified by our prediction model or our prediction had corrected the problem in the self-reported T2D by further investigating: 1) their follow-up time and current risk in the Cox regression model; 2) HbA1c and fasting glucose; 3) the accuracy of self-reported disease status.

**Figure 5.**
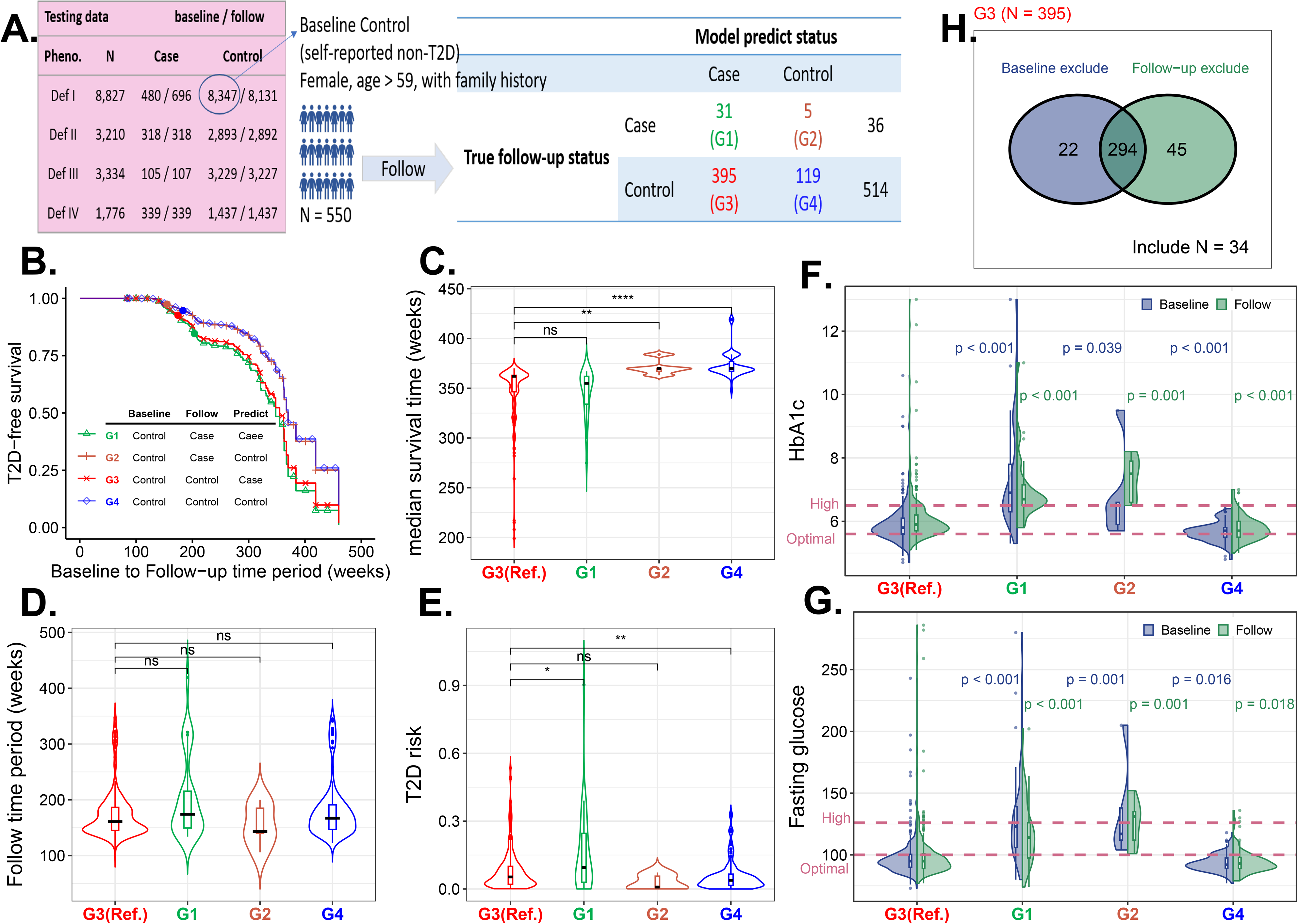
T2D early detection using our prediction model (Phenotype Definition IV; age, sex, family, and PRS). **(A) Four subgroups.** The individuals who are females aged >59 with a T2D family history and initially reported as non-T2D in the baseline are divided into four categories based on the T2D status predicted by our model and the self-reported T2D status in the follow-up. The four groups are G1 – True Positive (i.e., predicted as T2D and self-reported as T2D in the follow-up), G2 – False Negative (i.e., predicted as non-T2D but self-reported as T2D in the follow-up), G3 – False Positive (i.e., predicted as T2D but self-reported as non-T2D in the follow-up, and G4 – True Negative (i.e., predicted as non-T2D and self-reported as non-T2D in the follow-up). **(B) Survival rate.** The predicted survival proportion curves for each subgroup are displayed. G3 had a lower survival rate than G4 and higher than G1, indicating that most individuals in G3 are possibly pre-diabetic. **(C) Median survival time.** The distributions of median survival time for each subgroup are displayed. G3 had a shorter median survival time than G4 and longer than G1, suggesting that most individuals in G3 are possibly pre-diabetic. **(D) Follow-up time.** The distributions of the time period from the baseline to the follow-up for each subgroup are displayed. G3 had a follow-up time similar to G1 and G4. **(E) T2D risk**. The distributions of T2D risk at the follow-up for each subgroup are displayed. G3 had a higher T2D risk in the follow-up than G4 and lower than G1, suggesting that most individuals in G3 are possibly pre-diabetic. **(F) HbA1c.** The distributions of HbA1c at the baseline and follow-up for each subgroup are displayed. G3 had a higher HbA1c than G4 and lower than G1, indicating that most individuals in G3 are possibly pre-diabetic. **(G) Fasting glucose.** The distributions of fasting glucose at the baseline and follow-up are displayed. The results show that G3 had higher fasting glucose than G4 and lower than G1, indicating that most individuals in G3 are possibly pre-diabetic. **(H) Phenotype definition.** Many individuals in G3 cannot satisfy the T2D Phenotype Definition IV. Only 37 of 404 individuals passed the phenotype inclusion criteria. The self-reported T2D in G3 does not match the general definition of T2D based on HbA1C and fasting glucose, indicating that most individuals in G3 are possibly pre-diabetic.

First, compared to G4 (“true negative”), G3 had a significantly lower T2D-free probability (**Fig. 5B**), shorter median survival time (**Fig. 5C**), higher T2D-risk under similar follow-up time (**Fig. 5D** and **Fig. 5E**), higher HbA1c (**Fig. 5F**), and higher fasting glucose (**Fig. 5G**). Second, compared to G1 (“true positive”), G3 had a comparable survival rate (**Fig. 5B**), median survival time (**Fig. 5C**), and T2D-risk under similar follow-up time (**Fig. 5D** and **Fig. 5E**) but lower HbA1c (**Fig. 5F**) and fasting glucose (**Fig. 5G**). We didn’t compare G2 and G3 because of the small sample size in G2.

Finally, among the 395 participants in G3, 80.76% of them were removed from our previous analysis because their baseline HbA1c and fasting glucose violated the criteria for the phenotype definition (**Fig. 1C**); 339 participants were removed because of their follow-up HbA1c or fasting glucose violated the formal non-T2D criterion; only 34 self-reported non-T2D were really non-T2D participants who had HbA1C <6.5% and fasting glucose <126 mg/dL (**Fig. 5H**). Overall, the results consistently indicate that G3 represents individuals in a pre-T2D stage, which can be detected early by our model.

### Genetic-imaging integrative analysis

#### Model performance and essential features

The model that combined four types of image features performed best. Moreover, the model based on BMD image features exhibited a higher AUC, accuracy, specificity, and F1 than the models based on any other three types of images (**Fig. 6A**). The models based on image features had an AUC of 0.898 higher than the ones of genetic information (AUC = 0.677) and demographic factors (AUC = 0.843). Integrating image features, genetic information, and demographic factors increased AUC to 0.949 (**Fig. 6B**); the results for each of the four images are also provided (**Fig. S9)**. Accuracy, sensitivity, specificity, and F1 of the model in the first testing data were 0.871, 0.878, 0.870, and 0.663, respectively, based on a classification threshold 0.03. The model also performed reasonably well in the second testing dataset with AUC = 0.929, Accuracy = 0.854, Sensitivity = 0.789, Specificity = 0.862, and F1 = 0.558. According to the estimated feature importance in the best XGBoost model, all genetic factors (PRS), four types of medical images, and demographic variables provided informative features for risk assessment, such as PRS (genetics), family history and age (demographic factors), fatty liver (ABD images), end-diastolic velocity in the right common carotid artery (VAS images), RR interval (ECG images), and spine thickness (BMD images) (**Fig. 6C**).

**Figure 6.**
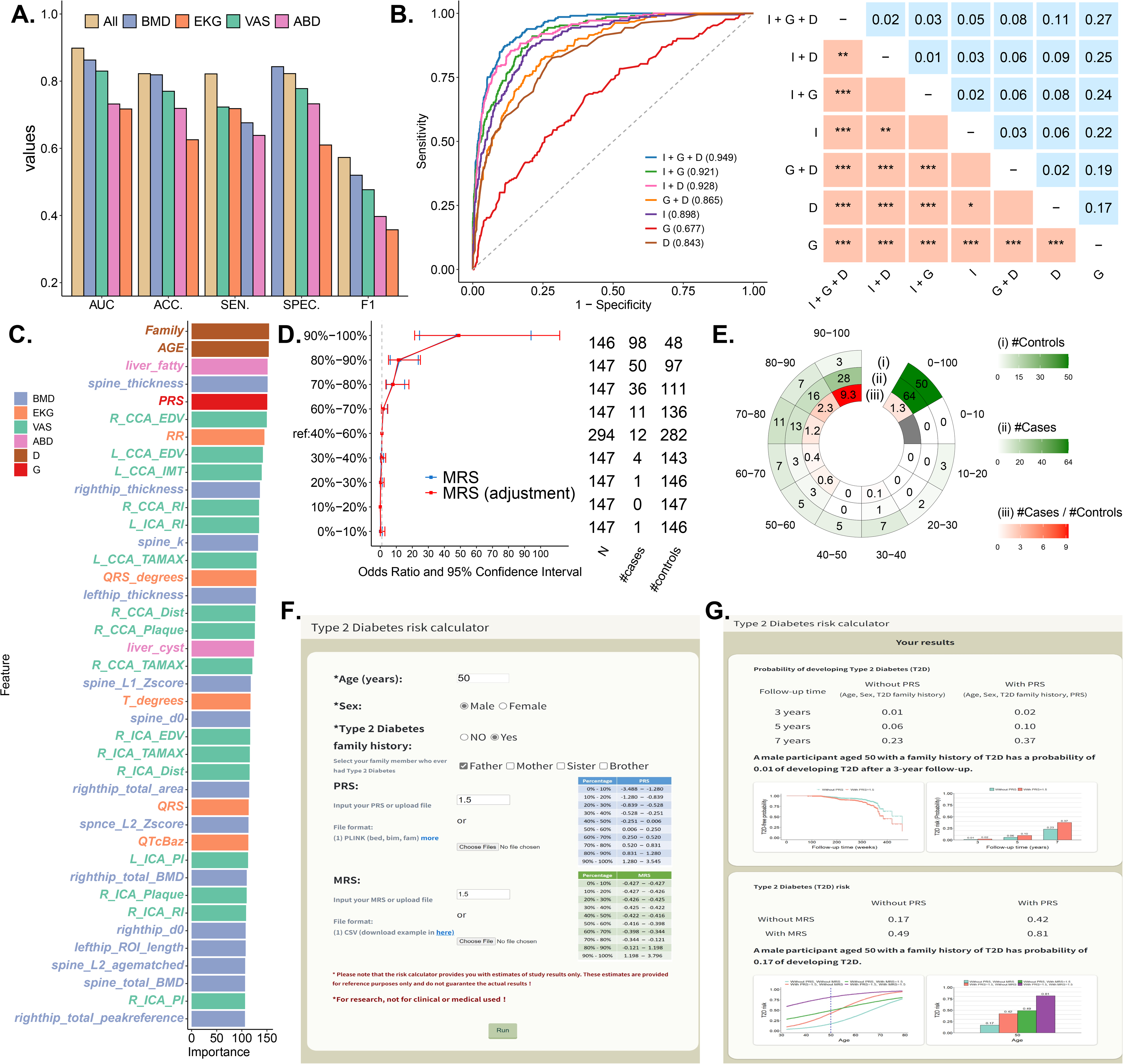
Results in the genetic-image integrative analysis. (**A**) Performance comparison of medical imaging data analysis. The area under the receiver operating characteristic (ROC) curve (AUC), accuracy (ACC), sensitivity (SEN), specificity (SPE), and F1 score are compared for the integrative analysis of four types of medical images (All) and individual medical imaging analyses, including BMD, EKG, VAS, and ABD. The analysis combining all four medical imaging data types shows the highest performance. **(B) The model that combines four types of medical imaging, PRS, and demographic variables shows the highest AUC of 0.949.** ROC plots and the corresponding AUC for the models considering medical imaging features (I), genetic PRS (G), and demographic variables, including age, sex, T2D family history (D), and their combinations. **(C) An optimal model combining medical imaging, PRS, and demographic variables.** The best model’s top 20 features with a high feature impact include the medical imaging, genetic, and demographic features. The high-impact image features include ABD-related features, including fatty liver and RR; BMD-related features, including spine and right hip thickness; EKG-related features, including RR and QRS degrees; and VAS-related features, including R CCA EDV and L CCA EDV. The high-impact genetic features: PRS. The high-impact demographic features: T2D family history and age. **(D) Positive correlation between MRS and T2D odds ratio.** In each decile of MRS based on four types of medical images, the odds ratio of T2D risk and its 95% confidence interval were calculated based on an unadjusted model (blue line) and an adjusted model considering age, sex, and T2D family history (red line), with the MRS group in the 40%–60% decile serving as the reference group. **(E) High-risk group.** A high-risk group was identified as males older than 59 with a T2D family history and falling into the PRS group in the 80%–100% decile. The figures from the inner to the outer in the chart display (i) the case-to-control ratio, (ii) the number of cases, and (iii) the number of controls in the MRS decile subgroups. **(F) Input page of the online T2D prediction website.** Personal information, including age, sex, family history of T2D, PRS, and MRS, is input to calculate T2D risk. PRS and MRS are optional, and a reference distribution is provided. **(G) Output page of the online T2D prediction website.** The results include the hazard of developing T2D in 3, 5, and 7 years for persons without and with inputted PRS data based on a Cox regression model. The probability of developing T2D without and with PRS and MRS based on a logistic regression model is also provided.

#### Multi-image risk score (MRS)

Each participant’s multi-image risk score (MRS) was calculated (see **Methods**). The odds ratio and its confidence interval for the association between MRS and T2D are shown by percentiles of MRS (**Fig. 6D**). Compared to the participants in the 40%–60% MRS decile group, the risk of T2D increased with MRS. Of importance, we further identified that, for the men older than 54 years old with a family history of T2D, the case vs. control ratio of sample size was 9.3 in the 90%–100% decile group, much higher than 1.3, which MRS was not considered (**Fig. 6E**).

### Online T2D-risk assessment

We have established a website for users to calculate their T2D risk online. To obtain the risk assessment, users need to provide age, sex, family history of relatives, PRS, and MRS (**Fig. 6F**). PRS and MRS can be entered manually or uploaded as a file (**Supplemental Text 2**). Additionally, we have provided PRS and MRS risk percentages based on the study population as a reference. The online risk assessment offers information, including the risk of developing T2D over 3, 5, and 7 years, T2D-free probability, and T2D risk with and without considering PRS (**Fig. 6G**). The assessment takes both PRS and MRS into account (**Fig. 6G**). For example, consider a 50-year-old male with a family history of T2D and PRS 1.5 and MRS 1.5. Without considering PRS, the risk (probability) of developing T2D after a 7-year follow-up is 0.23. However, when PRS is included, the risk increases to 0.37. Furthermore, considering MRS further increases the risk to 0.81. The online tool provides these valuable insights to users based on their input data.

## Discussions

In this study, we compared two prediction models based on GWAS data. The first model used SNPs from our GWAS, which had a limited sample size. The second model utilized SNPs from previously published GWASs with a considerably larger sample size. Interestingly, the latter approach resulted in a higher prediction AUC. Additionally, we constructed PRS using two methods. The first method involved significant SNPs from our own GWAS with a limited sample size, while the second method utilized summary statistics of whole-genome SNPs from GWASs with a considerably larger sample size. Notably, the latter approach resulted in a higher prediction AUC. These findings emphasize the significant influence of sample size in GWAS, PRS construction, and subsequent classification and prediction analyses, consistent with prior research ^47^. Consequently, in situations where the sample size is limited, we propose utilizing external genetic information such as SNPs and summary statistics from published studies with larger sample sizes, which not only facilitates the development of a more predictive PRS and model but also reduces computational overhead ^48^.

Our study emphasizes the superiority of disease family history as a predictor of T2D compared to T2D-associated SNPs and PRS. The inclusion of genetic factors such as significant SNPs and PRS as additional predictors, given family history, only results in modest improvements in the model’s predictive capability. Family history encompasses genetic and environmental influences, which are crucial in understanding the etiology of T2D ^49^. Additionally, we observed that the disease history of siblings provides more informative value for prediction than the disease history of parents ^50^.

T2D subgrouping can facilitate the implementation of precision medicine in clinical practice, particularly when utilizing complex data ^51^. This study demonstrated a positive association between PRS and MRS with T2D risk. Notably, we identified a high-risk subgroup of women older than 59 years with a family history of T2D, where the case vs. control ratio of sample size in the 80%–100% PRS decile group ranged from 7 to 13, significantly higher than the overall population. Similarly, for MRS, we found a high-risk subgroup of men older than 54 years with a family history of T2D, where the case vs. control ratio of sample size in the 90%–100% MRS decile group was 9.3, considerably higher than the ratio of 1.3 when MRS was not considered.

These results demonstrate the utility of PRS and MRS in identifying high-risk subgroups for T2D. In the PRS-CSx method, we considered three weighting methods to combine several population-specific PRSs into the final PRS: (1) an equal weight, (2) the population-specified weight, and (3) the meta-effect size for each SNP. Our results showed that the meta-effect size obtained a worse performance. The population-specified weight performed best; however, the result may vary between cohorts.

In this study, our PRS based on PRS-CSx achieved an AUC of 0.732 for T2D prediction. The AUC increased to 0.915 after further including age, sex, and family history of T2D into the prediction model. When comparing our results with the previous publications, Khera et al. ^52^ achieved an AUC of 0.725 using a logistic regression that included age, sex, and PRS constructed with LDpred ^53^. Imamura *et al.* ^14^ achieved an AUC of 0.648 with a PRS constructed by 49 T2D-associated SNPs with LD weights, and the AUC increased to 0.787 after including age, sex, and BMI. Ge *et al.* ^18^ achieved an AUC of 0.694 with a PRS constructed using summary statistics from three large-scale GWASs. Walford et al. ^54^ achieved an AUC of 0.641 with a PRS constructed by 63 SNPs, age, and sex. In summary, our study utilized phenotype refinement through HbA1c and fasting glucose, employed XGBoost with superior performance, and considered the family history of T2D as a critical factor for T2D prediction, leading to improved performance compared to previous studies.

Including environmental factors such as education level, drinking level, exercise habit, and the number of exercise types in our models increased prediction accuracy for non-T2D participants but decreased accuracy for T2D cases. The overall improvement in prediction performance achieved by including these environmental factors was relatively modest and did not reach statistical significance. Similarly, including SNP-SNP interactions in the models did not lead to a significant improvement. While SNP-SNP interactions have been proposed as a potential explanation for missing heritability ^55^, our findings indicate that incorporating these interactions does not provide additional benefits when PRS is already included in the model. This could be attributed to PRS already capturing a substantial portion of the genetic component, making incorporating SNP main effects and SNP-SNP interactions less impactful.

This study demonstrates good ability in detecting T2D cases, but we observed that some self-reported non-T2D individuals might be misclassified as T2D cases. Further investigation revealed that these cases represent individuals in a pre-T2D stage. Firstly, their T2D risk at the follow-up time was higher than true non-T2D participants but lower than the confirmed T2D cases, indicating an elevated but not fully developed risk. Secondly, these individuals exhibited higher HbA1c and fasting glucose levels than true non-T2D participants, albeit lower than confirmed T2D cases, suggesting a pre-T2D stage. Lastly, when redefining the phenotype using HbA1c and fasting glucose, a majority of these participants did not meet the inclusion criteria for the control group, further suggesting that they may not be truly non-T2D participants.

Considering these factors, it is evident that although these “misclassified” participants are self-reported as non-T2D, they are likely in a pre-T2D stage, with an increased risk of developing T2D in the future. It is crucial to follow up with these individuals, monitor their condition closely, and implement preventive interventions to mitigate the risk of T2D development.

The integration of genetics and medical imaging data into risk assessment shows excellent potential for enabling early T2D detection and prevention, albeit at a higher cost. Practical examples from health examinations and screenings, such as the MJ Health Survey Database ^56^, provide compelling evidence for successfully incorporating these data into real-world practices. These examples highlight the valuable role that genetics and medical imaging data can play in enhancing risk assessment and underscore the potential benefits of integrating these approaches for improved disease management and prevention.

Another limitation of our study is that due to limited follow-up time in the TWB, only a limited number of participants experienced a change in T2D status from baseline to follow-up, particularly for redefining the phenotype using HbA1c and fasting glucose. To assess the early detection capability of our model for T2D, we are currently addressing this issue by monitoring the participants who exhibited changes in self-reported T2D status from baseline to follow-up in our Cox regression model. This limitation can be overcome in future studies as the TWB continues to track these samples. In addition, conducting a cohort survey or clinical trial is warranted to evaluate the high-risk subgroups identified by our PRS and MRS for future precision T2D medicine.

## Conclusion

In conclusion, our study surpassed previous research in the prediction and classification of T2D. We successfully developed artificial intelligence models that effectively combined genetic markers, medical imaging features, and demographic variables for early detection and risk assessment of T2D. PRS and MRS were instrumental in identifying high-risk subgroups for T2D risk assessment. To facilitate online T2D risk evaluation, we have also created a dedicated website.

## Data and code availability

The data analyzed in this study were obtained from the Taiwan Biobank with proper approval. As the data are subject to ownership rights held by the Taiwan Biobank, they have not been deposited in a public repository. Researchers interested in accessing the data must do so through a formal application process, subject to approval by the Taiwan Biobank. Detailed instructions on requesting data access can be found on the Taiwan Biobank’s official website (https://www.twbiobank.org.tw/index.php). The codes developed in this study are available upon request.

## Supplemental information

Supplemental information is online available.

## Data Availability

All data produced in the present work are contained in the manuscript

https://hcyang.stat.sinica.edu.tw/software/T2D_web/header.php

## Acknowledgments

This work was supported by research grants from the Academia Sinica (AS-PH-109-01 and AS-SH-112-01). Data application and use were approved by the Taiwan Biobank and the Institute Review Board (AS-IRB01-17049 and AS-IRB01-21009). We gratefully acknowledge the Taiwan Biobank for providing the data used in this research. We also extend our thanks to all the participants of the Taiwan Biobank for their invaluable contributions. Technical support in genotyping from the National Center for Genome Medicine of Taiwan is also acknowledged.

## Author contributions

H.C.Y. conceptualized and supervised the study. Y.J.H. performed data curation and applied software. Y.J.H. & H.C.Y. conducted formal data analysis, result visualization, and wrote the paper. C.h.C. & H.C.Y. provided funding acquisition and resources. H.C.Y., Y.J.H., and C.h.C. validated the results.

## Web resources

An online risk assessment website for T2D is available at https://hcyang.stat.sinica.edu.tw/software/T2D_web/header.php

