## Supplemental files for "AI-Enhanced Integration of Genetic and Medical Imaging Data for Risk Assessment of Type 2 Diabetes"

Supplemental information of the paper entitled "AI-Enhanced Integration of Genetic and Medical Imaging Data for Risk Assessment of Type 2 Diabetes"

Yi-Jia Huang[1], Chun-houh Chen[2], and Hsin-Chou Yang[1,2,]*

[1]Institute of Public Health, National Yang-Ming Chiao-Tung University, Taipei, Taiwan
[2]Institute of Statistical Science, Academia Sinica, Taipei, Taiwan

*Corresponding author: Hsin-Chou Yang, Institute of Statistical Science, Academia Sinica. No. 128, Sec. 2, Academia Road, Nankang 115, Taipei, Taiwan

(Fax) 886-2-27886833

(Tel) 886-2-27875686

### Supplemental Information


| | | |
|---|---|---|
| Figure S1 | Sample sizes in the two analyses in this study................................ | 2 |
| Figure S2 | Performance of the SNP predictors selected from published GWAS or our GWAS with different p-value thresholds.................. | 3 |
| Figure S3 | The T2D family history's performance combined with other demographic and genetic predictors............................................ | 4 |
| Figure S4 | Manhattan plot of genome-wide association study...................... | 5 |
| Figure S5 | Prediction AUCs of all the models................................................. | 6 |
| Figure S6 | Classification AUCs of all the models............................................ | 7 |
| Figure S7 | Risk factors for T2D....................................................................... | 8 |
| Figure S8 | Effect of doing exercise on HbA1c................................................. | 9 |
| Figure S9 | ROC plots and the corresponding AUC for the models considering the combinations of image report variables, genetic factors, and demographic factors................................................ | 10 |
| Table S1 | Description of image report features............................................ | 11 |
| Table S2 | Data used in the two analyses in this study................................... | 16 |
| Table S3 | Performance evaluation for the models that add environmental factors or SNP x SNP interactions as predictors............................ | 17 |
| Supp Text 1 | Sources of genetic variable data.................................................... | 18 |
| Supp Text 2 | Input data for an online T2D-risk assessment................................ | 19 |
| References | ............................................................................................. | 20 |

#### Supplementary Figures

Total number of participants: N = 68,911 (= 50,984 + 9,763 + 8,164)

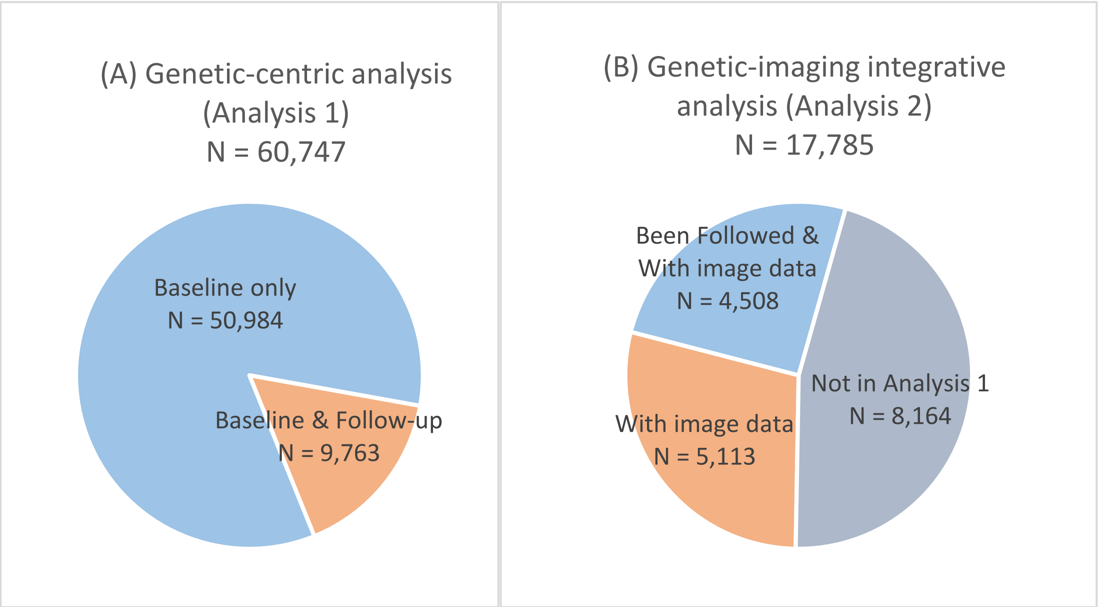


**Figure S1. Sample sizes in the two analyses in this study. (A) Analysis 1: Genetic-centric analysis.** The genetic-centric analysis (**Analysis 1**) included 60,747 participants, including 50,984 participants with only baseline data and 9,763 participants with both baseline and follow-up data. **(B) Analysis 2: Genetic-imaging integrative analysis.** The genetic-imaging integrative analysis (**Analysis 2**) included a total of 17,785 participants who had both baseline and follow-up data, of which 9,198 (= 4,085 + 5,113) participants were included in **Analysis 1,** and additional 8,164 participants were only included in **Analysis 2**. The unique participants in the two analyses in this study were 68,911 (= 50,984 + 9,763 + 8,164).

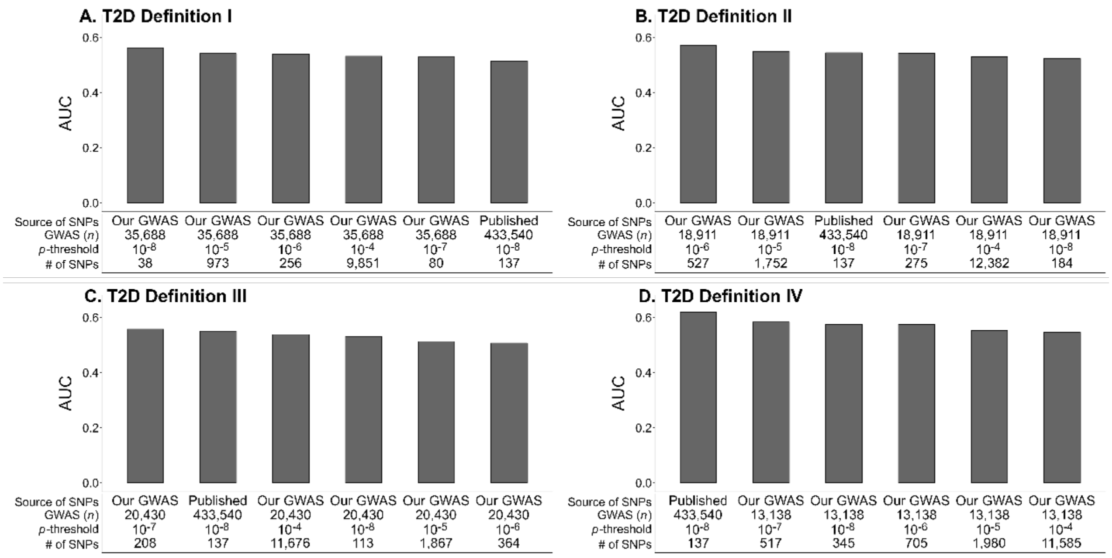


**Figure S2. Performance of the SNP predictors selected from published GWAS or our GWAS with different p-value thresholds.** AUC is displayed via a bar chart. AUCs of different SNPs are listed from high to low. (**A**) T2D Definition I; (**B**) T2D Definition II; (**C**) T2D Definition III; (**D**) T2D Definition IV. In general, the difference in AUCs is limited.

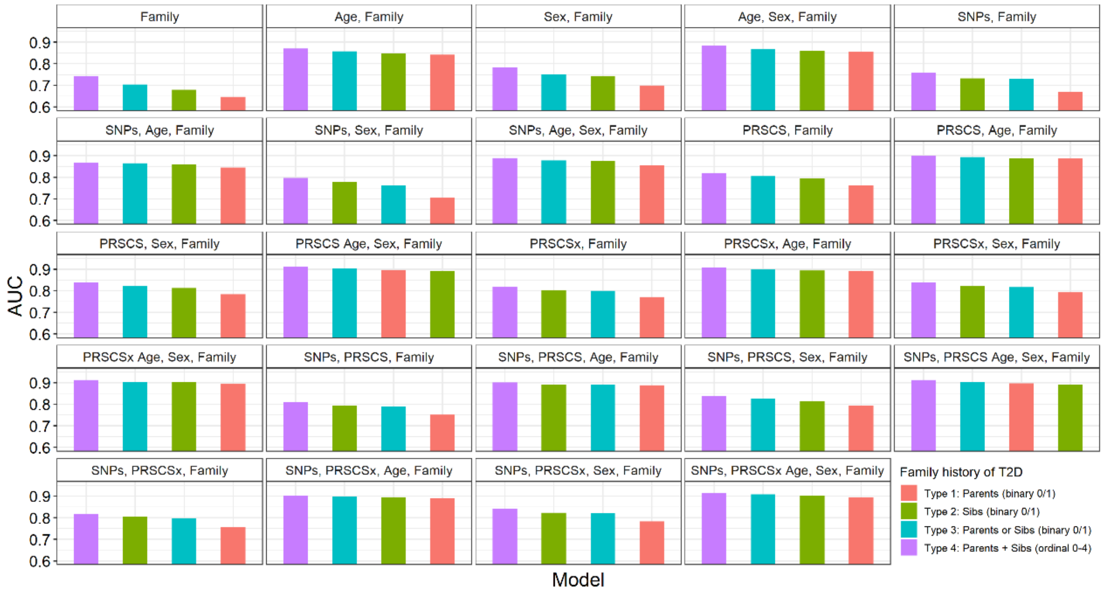


**Figure S3. The T2D family history's performance combined with other demographic and genetic predictors.** The model considering both parents' and sibs' T2D history (purple bars) outperforms other types of T2D family history.

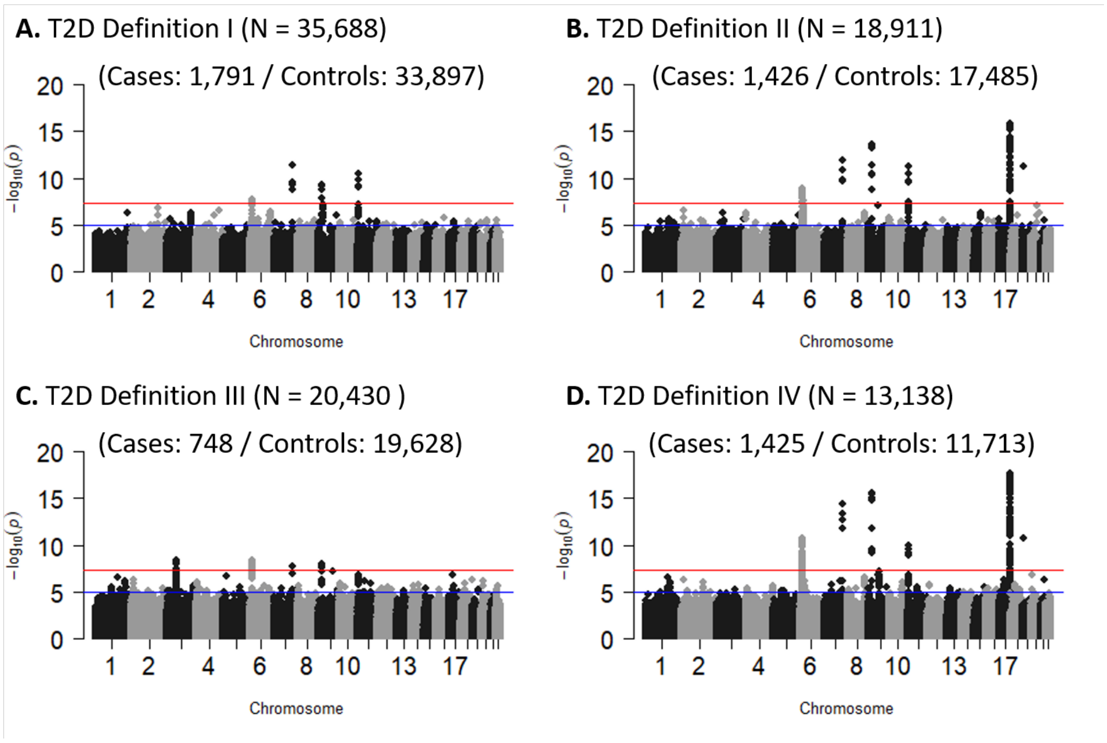


**Figure S4. Manhattan plot of genome-wide association study.** T2D-associated SNPs, HbA1c, and fasting glucose were identified. (**A**) T2D Definition I; (**B**) T2D Definition II; (**C**) T2D Definition III; (**D**) T2D Definition IV.

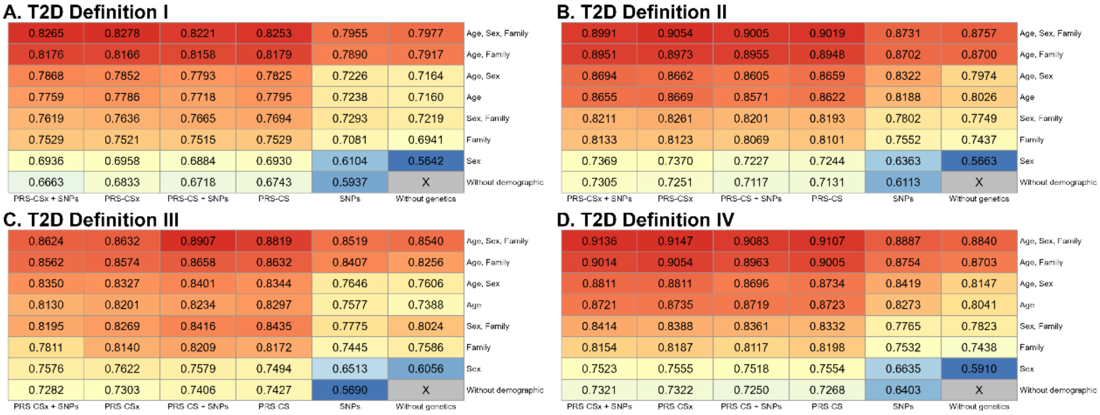


**Figure S3. Prediction AUCs of all the models.** A heatmap shows the models' prediction AUCs in four T2D definitions. The genetic variables are shown on the X-axis, and the demographic variables are shown on the Y-axis. **(A)** T2D Definition I; **(B)** T2D Definition II; **(C)** T2D Definition III; **(D)** T2D Definition IV.

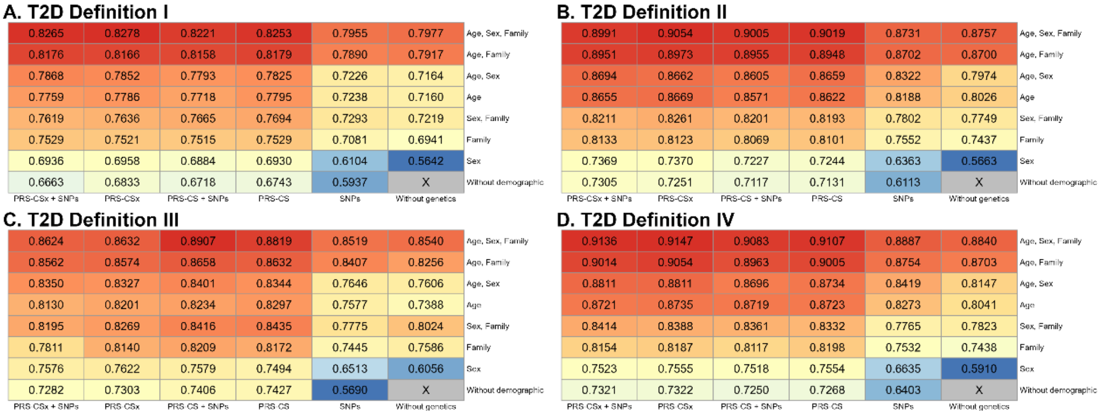


**Figure S4. Classification AUCs of all the models.** A heatmap shows the models' classification AUCs in four T2D definitions. The genetic variables are shown on the X-axis, and the demographic variables are shown on the Y-axis. **(A)** T2D Definition I; **(B)** T2D Definition II; **(C)** T2D Definition III; **(D)** T2D Definition IV.

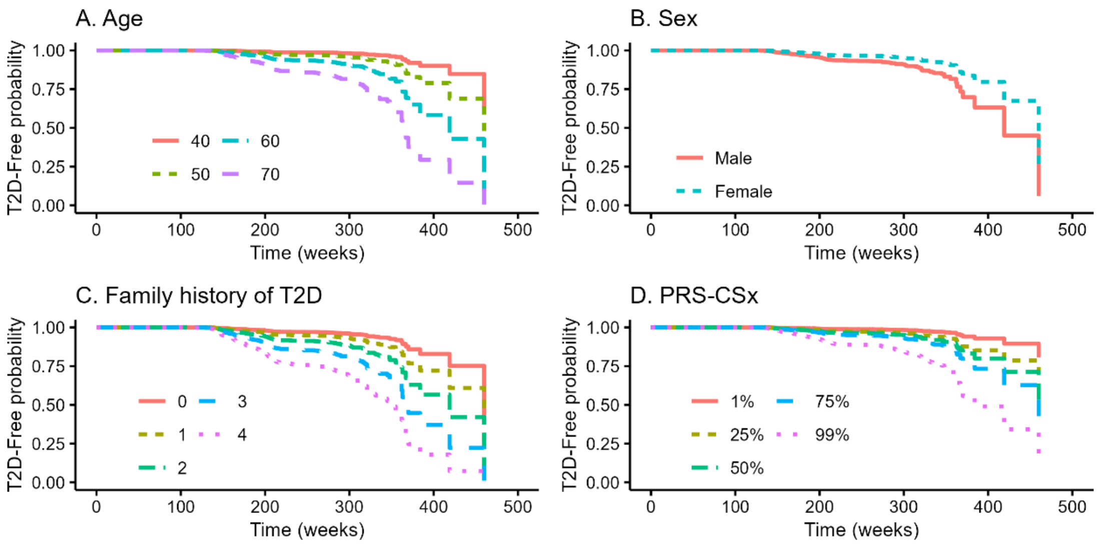


**Figure S5. Risk factors for T2D.** Kaplan-Meier curves reveal significant T2D risk factors (high-risk level). (**A**) Age (older persons); (**B**) Sex (males); (**C**) T2D family history (the number of parents and siblings who had T2D); and (**D**) PRS (high decile PRS group).

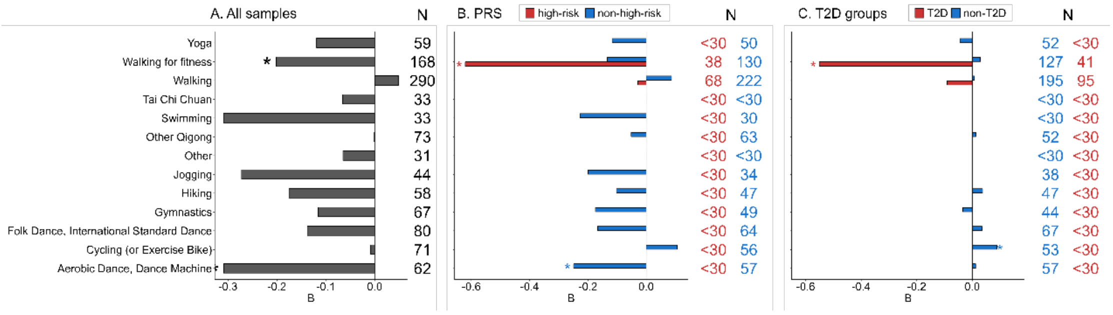


**Figure S6. Effect of doing exercise on HbA1c.** Beta coefficients (*B*) for various types of exercise are displayed via a bar chart. The figures on the right-hand side indicate the number of individuals for each type of exercise. Only the result of an analysis containing a sample size > 30 is shown. **(A) All samples**; **(B) PRS.** Beta coefficients for the high-risk group (red bar) and non-high-risk group (blue) are displayed; **(C) T2D groups.** Beta coefficients for the T2D group (red bar) and non-T2D group (blue) are displayed. Walking for fitness was found to be significantly negatively associated with T2D in all samples, the high-PRS group and the T2D group.

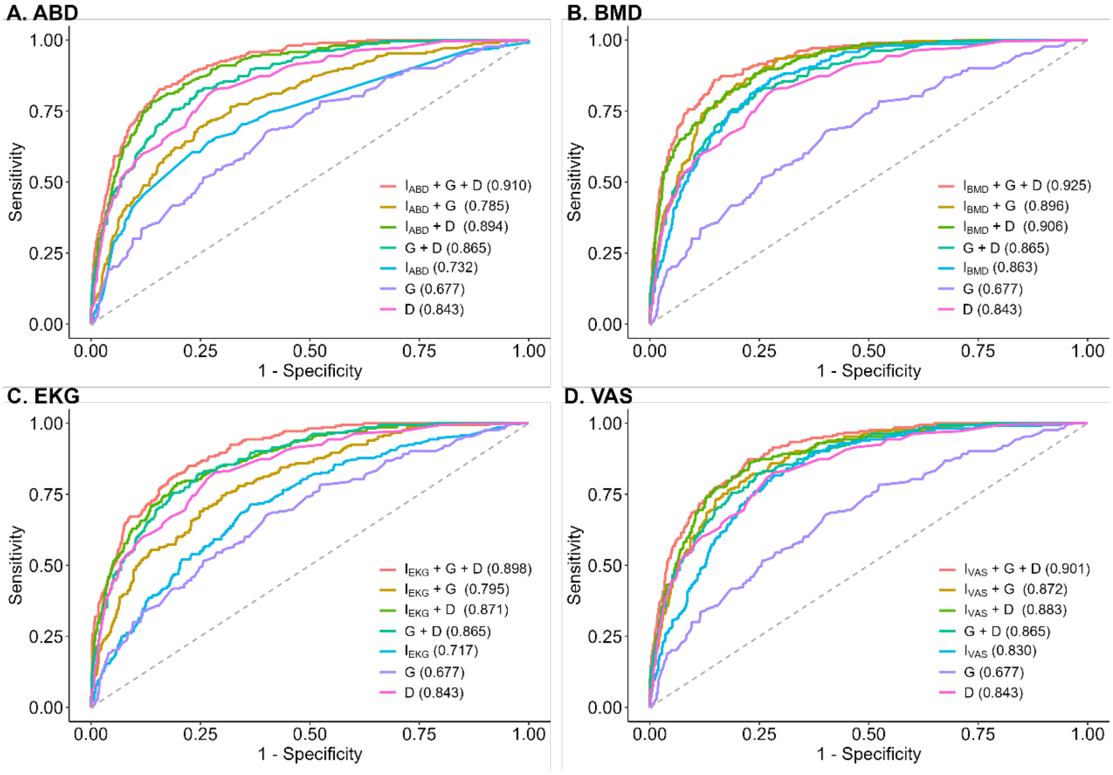


**Figure S7. ROC plots and the corresponding AUC for the models considering** the **combination of image report variables, genetic factors, and demographic factors.** ROC plots and the corresponding AUC for the models considering medical image features (I), genetic PRS (G), and demographic variables, including age, sex, T2D family history (D), and their combinations. **(A)** ABD features; **(B)** BMD features; **(C)** EKG features; **(D)** VAS features. BMD had the best performance.

#### Supplementary Tables

**Table S1. Description of image report features.**

| Variable | Description |
|---|---|
| | ABD |
| liver_tumor | liver tumor |
| liver_cyst | liver cyst |
| liver_cal | liver calcification |
| liver_hemangioma | liver hemangioma |
| liver_fibrosis | liver fibrosis |
| liver_chronic | Chronic liver |
| liver_fatty | Fatty liver |
| GB_polyp | Gallbladder polyps |
| GB_stone | Gallbladder stone |
| CBD_dilatation | Common bile duct dilatation |
| CBD_stone | Common bile duct stone |
| pancreas_cyst | Pancreatic cyst |
| pancreas_tumor | Pancreatic tumor |
| pancreas_cal | Pancreatic calcification |
| pancreas_stone | Pancreatic calcification |
| pancreas_dilatation | Pancreatic dilatation |
| pancreas_pancreatitis | Pancreatitis |
| spleen_tumor | Spleen tumor |
| spleen_cyst | Spleen cyst |
| spleen_cal | Spleen calcification |
| spleen_accessory | Spleen accessory |
| spleen_splenomegaly | Spleen splenomegaly |
| kidney_cyst | Kidney cyst |
| kidney_stone | Kidney stone |
| kidney_cal | Kidney calcification |
| kidney_hydronephrosis | Kidney hydronephrosis |
| kidney_polycystic | Kidney polycystic |
| kidney_tumor | Kidney tumor |
| | VAS |
| R_CCA_Plaque | Plaque in the right common carotid artery |
| R_CCA_PSV | Peak systolic velocity in the right common carotid artery |

**Table S1 (continued).**

| Variable | Description |
|---|---|
| | VAS |
| R_CCA_EDV | End-diastolic velocity in the right common carotid artery |
| R_CCA_EDV | End-diastolic velocity in the right common carotid artery |
| R_CCA_TAMAX | Time average maximum mean velocity of right common carotid artery |
| R_CCA_PI | Pulsatility Index in the right common carotid artery |
| R_CCA_RI | Resistive index in the right common carotid artery |
| R_CCA_Dist | Diameter of the right common carotid artery |
| R_CCA_IMT | Intima-media thickness in the right common carotid artery |
| R_ICA_Plaque | Plaque in the right internal carotid artery |
| R_ICA_PSV | Peak systolic velocity in the right internal carotid artery |
| R_ICA_EDV | End-diastolic velocity in the right internal carotid artery |
| R_ICA_TAMAX | Time average maximum mean velocity of right internal carotid artery |
| R_ICA_PI | Pulsatility Index in the right internal carotid artery |
| R_ICA_RI | Resistive index in the right internal carotid artery |
| R_ICA_Dist | Diameter of the right internal carotid artery |
| L_CCA_PSV | Peak systolic velocity in the left common carotid artery |
| L_CCA_EDV | End-diastolic velocity in the left common carotid artery |
| L_CCA_TAMAX | Time average maximum mean velocity of left common carotid artery |
| L_CCA_PI | Pulsatility Index in the left common carotid artery |
| L_CCA_RI | Resistive index in the left common carotid artery |
| L_CCA_Dist | Diameter of the left common carotid artery |
| L_CCA_IMT | Intima-media thickness in the left common carotid artery |
| L_ICA_Plaque | Plaque in the left common carotid artery |
| L_ICA_PSV | Peak systolic velocity in the left internal carotid artery |

**Table S1 (continued).**

| Variable | Description |
|---|---|
| | VAS |
| L_ICA_EDV | End-diastolic velocity in the left internal carotid artery |
| L_ICA_TAMAX | Time average maximum mean velocity of left internal carotid artery |
| L_ICA_PI | Pulsatility Index in the left internal carotid artery |
| L_ICA_RI | Resistive index in the left internal carotid artery |
| L_ICA_Dist | Diameter of the left internal carotid artery |
| | EKG |
| QRS | QRS complex |
| QTcBaz | Corrected QT interval (Bazett's formula) |
| PR | PR interval |
| P | P wave |
| RR | RR interval |
| PP | PP interval |
| P_degrees | P-wave axis |
| QRS_degrees | QRS axis |
| T_degrees | T-wave axis |
| ECG_summary | ECG summary |
| | BMD |
| spine_k | Spine k |
| spine_d0 | Spine d0 |
| spine_thickness | Spine thickness |
| spine_ROI | Width of the region of interest in the spine |
| spine_ROI_length | Length of the region of interest in the spine |
| spine_L1_area | bone area in the lumbar spine (L1) |
| spine_L1_BMC | bone mineral content in the lumbar spine (L1) |
| spine_L1_BMD | bone mineral density in the lumbar spine (L1) |
| spine_L1_Tscore | lumbar spine (L1) T-Score |
| spine_L1_Zscore | lumbar spine (L1) Z-Score |
| spine_L1_peakreference | The peak reference value for the lumbar spine (L1) |
| spine_L1_agematched | Age-Matched value for lumbar spine (L1) |
| spine_L2_area | bone area in the lumbar spine (L2) |
| spine_L2_BMC | bone mineral content in the lumbar spine (L2) |
| spine_L2_BMD | bone mineral density in the lumbar spine (L2) |
| spine_L2_Tscore | lumbar spine (L2) T-Score |

**Table S1 (continued).**

| Variable | Description |
|---|---|
| | BMD |
| spnce_L2_Zscore | lumbar spine (L2) Z-Score |
| spine_L2_peakreference | The peak reference value for the lumbar spine (L2) |
| spine_L2_agematched | Age-Matched value for lumbar spine (L2) |
| spine_L3_area | bone area in the lumbar spine (L3) |
| spine_L3_BMC | bone mineral content in the lumbar spine (L3) |
| spine_L3_BMD | bone mineral density in the lumbar spine (L3) |
| spine_L3_Tscore | lumbar spine (L3) T-Score |
| spine_L3_Zscore | lumbar spine (L3) Z-Score |
| spine_L3_peakreference | The peak reference value for the lumbar spine (L3) |
| spine_L3_agematched | Age-Matched value for lumbar spine (L3) |
| spine_L4_area | bone area in the lumbar spine (L4) |
| spine_L4_BMC | bone mineral content in the lumbar spine (L4) |
| spine_L4_BMD | bone mineral density in the lumbar spine (L4) |
| spine_L4_Tscore | lumbar spine (L4) T-Score |
| spine_L4_Zscore | lumbar spine (L4) Z-Score |
| spine_L4_peakreference | The peak reference value for the lumbar spine (L4) |
| spine_L4_agematched | Age-Matched value for lumbar spine (L4) |
| spine_total_area | Spine total area |
| spine_total_BMC | Spine total bone mineral content |
| spine_total_BMD | Spine total bone mineral density |
| spine_total_Tscore | Spine total T-Score |
| spine_total_Zscore | Spine total Z-Score |
| spine_total_peakreference | Spine total peak reference value |
| spine_total_agematched | Spine total age-matched value |
| lefthip_K | Left hip k |
| lefthip_d0 | Left hip d0 |
| lefthip_thickness | The thickness of the left hip |
| lefthip_ROI_width | Width of the region of interest in the left hip |
| lefthip_ROI_length | Length of the region of interest in the left hip |
| lefthip_neck_area | The neck area of the left hip |
| lefthip_neck_BMC | Neckbone mineral of the left hip |
| lefthip_neck_BMD | Neckbone mineral density of the left hip |
| lefthip_neck_Tscore | Neck T-Score of the left hip |
| lefthip_neck_Zscore | Neck Z-Score of the left hip |
| lefthip_neck_peakreference | Neck peak reference value of the left hip |

**Table S1 (continued).**

| Variable | Description |
|---|---|
| | BMD |
| lefthip_neck_agematched | Neck age-matched value of the left hip |
| lefthip_total_area | Neck total area of the left hip |
| lefthip_total_BMC | Neck total bone mineral of the left hip |
| lefthip_total_BMD | Neck total bone mineral density of the left hip |
| lefthip_total_Tscore | Neck total T-Score of the left hip |
| lefthip_total_Zscore | Neck total Z-Score of the left hip |
| lefthip_total_peakreference | Neck total peak reference value of the left hip |
| lefthip_total_agematched | Neck total age-matched value of the left hip |
| righthip_K | Right hip k |
| righthip_d0 | Right hip d0 |
| righthip_thickness | The thickness of the right hip |
| righthip_ROI_thickness | Width of the region of interest in the right hip |
| righthip_ROI_length | Length of the region of interest in the right hip |
| righthip_neck_width | Neck width of the right hip |
| righthip_neck_area | The neck area of the right hip |
| righthip_neck_BMC | Neckbone mineral of the Left Hip |
| righthip_neck_BMD | Neckbone mineral density of the right hip |
| righthip_neck_Tscore | Neck T-Score of the right hip |
| righthip_neck_Zscore | Neck Z-Score of the right hip |
| righthip_neck_peakreference | Neck peak reference value of the right hip |
| righthip_neck_agematched | Neck age-matched value of the right hip |
| righthip_total_area | Neck total area of the right hip |
| righthip_total_BMC | Neck total bone mineral of the right hip |
| righthip_total_BMD | Neck total bone mineral density of the right hip |
| righthip_total_Tscore | Neck total T-Score of the right hip |
| righthip_total_Zscore | Neck total Z-Score of the right hip |
| righthip_total_peakreference | Neck total peak reference value of the right hip |
| righthip_total_agematched | Neck total age-matched value of the right hip |
| MEASURE_BONE_POSE | Dominant limb for bone density measurement |
| BONE_EXAM_RESULT | Bone Stiffness Index |
| YOUNG_ADULT | Percentile in young adults |
| T_SCORE | T-SCORE |
| AGE_MATCHED | Percentile in age-matched adults |
| Z_SCORE | Z-SCORE |

| | Baseline | Follow-up (①Early / ②Late) |
|---|---|---|
| **TWB Data** | Genotyping data (TWB1.0, TWB2.0)<br>Questionnaire (Self-reported disease, Demo., Family history, Enviro.)<br>Blood and urine tests (HbA1c, GLU-AC) | ① / ② Medical images (ABD, BMD, EKG, VAS, TU)<br>① / ② Questionnaire (Self-reported disease, Demo., Family history, Enviro.)<br>① / ② Blood and urine tests (HbA1c, GLU-AC) |

**Analysis 1 – Genetic-centric analysis**

| | | | Baseline | Follow-up | Sample size |
|---|---|---|---|---|---|
| Classification | Mode building | Phenotype | Self-reported T2D status<br>HbA1c, GLU-AC | | 50,984 |
| | | Predictors | TWB2.0 imputed<br>Demo., Family history, Enviro. | | |
| | Mode testing | Phenotype | Self-reported T2D status<br>HbA1c, GLU-AC | | 8,827 |
| | | Predictors | TWB2.0 imputed<br>Demo., Family history, Enviro. | | |
| Prediction | Mode building | Phenotype | Self-reported T2D status<br>HbA1c, GLU-AC | | 50,984 |
| | | Predictors | TWB2.0 imputed<br>Demo., Family history, Enviro. | | |
| | Mode testing | Phenotype | | ① Self-reported T2D status<br>① HbA1c, GLU-AC | 8,827 |
| | | Predictors | TWB2.0 imputed<br>Demo., Family history, Enviro. | | |
| | Independent testing | Phenotype | | ② Self-reported T2D status<br>② HbA1c, GLU-AC | 936 |
| | | Predictors | TWB2.0 imputed<br>Demo., Family history, Enviro. | | |

**Analysis 2 – Genetic-imaging integrative analysis**

| | | | Baseline | Follow-up | Sample size |
|---|---|---|---|---|---|
| Classification | Mode building | Phenotype | | ① Self-reported T2D status<br>① HbA1c, GLU-AC | 5,864 |
| | | Predictors | TWB2.0 imputed | ① Demo., Family history, Enviro. | |
| | Mode testing | Phenotype | | ① Self-reported T2D status<br>① HbA1c, GLU-AC | 1,469 |
| | | Predictors | TWB2.0 imputed | ① Demo., Family history, Enviro. | |
| | Independent testing | Phenotype | | ② Self-reported T2D status<br>② HbA1c, GLU-AC | 444 |
| | | Predictors | TWB2.0 imputed | ② Demo., Family history, Enviro. | |

**Table S2. Data used in the two analyses in this study.** TWB data comprise data in the baseline and the follow-up. In the genetic-centric analysis (Analysis 1), classification models were built and tested based on the variables and phenotype data in the baseline. Prediction models were built based on the variables and phenotype data in the baseline. The model was tested based on the prediction variables in the baseline and T2D phenotype in the follow-up. In the genetic-imaging integrative analysis (Analysis 2), classification models were built and tested based on the predictors and phenotype data in the follow-up and further replicated based on the second independent testing dataset.

**Table S3. Performance evaluation for the models that add environmental factors or SNP x SNP interactions as predictors.** Demographic factors (Demo.) include age, sex, and family history of T2D. Genetic information includes PRS-CSx, SNP Main effect, and SNP interactions. Environmental factors include education, drinking experience, exercise habits, and the number of exercise types. SNP-SNP interactions include 1,059 SNP pairs ($p < 10^{-10}$). Moreover, SNP's main effect includes 1,191 SNPs from 1,059 pairs of SNPs.

| | Features | | | | | Performance | | | | |
|---|---|---|---|---|---|---|---|---|---|---|
| | Demo. | PRS-CSx | Environmental | SNPs Main effect | SNPs Interaction | AUC | Accuracy | Sensitivity | Specificity | F1 |
| Model 1 | O | O | X | X | X | 0.9147 | 0.8429 | 0.8437 | 0.8427 | 0.6722 |
| Model 2 | O | O | O | X | X | 0.9146 | 0.8440 | 0.8142 | 0.8511 | 0.6659 |
| Model 3 | O | O | X | X | O | 0.9111 | 0.7928 | 0.8879 | 0.7704 | 0.6206 |
| Model 4 | O | O | O | X | O | 0.9125 | 0.7675 | 0.9204 | 0.7314 | 0.6017 |
| Model 5 | O | O | X | O | O | 0.9044 | 0.7973 | 0.8702 | 0.7801 | 0.6211 |
| Model 6 | O | O | O | O | O | 0.9112 | 0.7866 | 0.9027 | 0.7592 | 0.6176 |

#### Supplementary Texts

**Supplemental Text 1: Sources of genetic variable data.**

Sources of genetic variable data were considered and compared (**Fig. 1C** and **Fig. 2B**): (i) a set of SNPs selected from our GWAS; (ii) a set of highly significant T2D-associated SNPs from publications; (iii) polygenic risk score (PRS) based on the GWAS summary statistics of T2D from publications; (iv) SNP-SNP and SNP-environment interactions. They are elaborated as follows.

As to (i), GWASs for four phenotypes (**Figs. 1B** and **1C**) were conducted using logistic regression with an adjustment for age, sex, and principal components PC1 to PC10 and an additive model of SNP. For each phenotype definition, different p-value thresholds ($p < 10^{-4}, 10^{-5}, 10^{-6}, 10^{-7}, 10^{-8}$) were applied.

As to (ii), a set of 137 highly significant T2D-associated SNPs from the Asian Genetic Epidemiology Network (AGEN) (Spracklen et al., 2020) that is a meta-analysis based on 77,414 T2D cases and 256,122 healthy controls from 23 GWASs.

As to (iii), SNP effects were estimated by using PRS-CS (Ge et al., 2019) based on the meta-GWAS summary statistics of T2D in East Asia in the DIAGRAM Consortium (Mahajan et al., 2022) and the linkage disequilibrium (LD) reference from the EAS population of the 1000 Genomes Project (Auton et al., 2015). PRS was calculated using PLINK (--score command) based on our genotype data, and 884,327 SNP effects were estimated using PRS-CS. Normalized PRS was standardized to mean = 0 and standard deviation = 1.

As to (iv), SNP effects were estimated by using PRS-CSx (Ruan et al., 2022) based on the meta-GWAS summary statistics of T2D in multiple populations, including (a) East Asian of 56,268 cases and 227,155 controls in the DIAGRAM Consortium (Mahajan et al., 2022); (b) European of 80,154 cases and 853,816 controls in the DIAGRAM Consortium (Mahajan et al., 2022); (c) South Asian of 16,540 cases and 32,952 controls in the DIAGRAM Consortium (Mahajan et al., 2022), and the LD reference from each of the three populations (EAS, EUR, and SAS). 884,327, 880,098, and 900,047 SNPs for EAS, EUR, and SAS were applied to our data to calculate the population-specific PRS for each individual using the PLINK (--score command). We combined the three population-specific PRS with equal weight to calculate a final PRS. R language was used to standardize the PRS to mean = 0 and standard deviation = 1.

(v) Finally, we considered all pairwise SNP-SNP and SNP-environment interactions. SNP–SNP interactions considered all pairwise combinations of SNPs, then selected SNP-SNP interaction pairs satisfying $p < 10^{-10}$ using PLINK --fast-epistasis command.

**Supplemental Text 2: Input data for an online T2D-risk assessment.**

As to the individual genotype files, users must upload their files following PLINK format containing bed, bim, and fam. Click “example” to download the example files. Users should upload their files to the medical imaging information following the CSV format. Click “example” to download an example file.
